# Expanding Risks: Medicaid Expansion and Data Security

**DOI:** 10.1101/2024.06.30.24309745

**Authors:** Jeffrey Clement, Brad N Greenwood, John D’Arcy, Corey Angst

**Author notes:** The authors thank Joshua Wright for his feedback during the development of this manuscript.

## Abstract

The Patient Protection and Affordable Care Act of 2010 led to the largest expansion of healthcare coverage since the instantiation of Medicare and Medicaid in 1965. Yet, limited attention has been given to the security aftereffects of the statute, specifically the potential for malfeasance in the form of consumer fraud and identity theft resulting from the vast influx of new patient data residing in various and highly dispersed sources. In this work, we fill this gap by exploiting the phased expansion of Medicaid into different states at different times. Using a difference in difference approach, we explore the data security-related aftereffects of the law. Results indicate a significant *decrease* in claims of consumer fraud after the expansion of Medicaid, with no robust effect on identity theft. In empirical extensions, we find a material drop in data breaches and compromised records after the expansion of Medicaid. Taken in sum, these findings suggest that the expansion of Medicaid had a consequential effect on the security of consumer data and created significant positive externalities for consumers.

---

The management and provision of healthcare in the United States remains one of the most divisive topics among its citizens. Emblematic of the heated debate is the continued battle over former President Obama’s signature domestic policy program, the Patient Protection and Affordable Care Act (ACA), also known as “Obamacare”, the constitutionality of which has been argued in front of the Supreme Court four separate times and which holds the distinction of being the most challenged statute in the nation’s history (1, 2).

Interestingly, despite the scrutiny afforded to the healthcare-related aspects of the statute, its societal implications beyond healthcare have received circumspect empirical consideration (with some notable exceptions (3, 4)). Indeed, while researchers point to increased vaccination rates, improved medical coverage among the nation’s youth, and coverage in underserved communities (5-7), as well as distinct differences in how young people care for themselves, little attention has been paid to its wider implications. This is striking given the aftereffects that traditionally spill from such a large injection of capital into dispersed and heterogeneous markets. In this work, we begin to close this gap by investigating an outcome that often characterizes expansive government spending: moral hazard and the fraud that accompanies it (8-11). We do so by investigating the effect of Medicaid expansion on rates of consumer fraud and identity theft.

The theoretical relationship between Medicaid expansion, consumer fraud, and identity theft is deeply murky. On the one hand, the conventional view would be that fraud and identity theft will likely rise with the expansion. To the extent that the expansion of Medicaid brought with it tremendous increases in coverage, the creation of individual health insurance markets, and an intense digitization of records (12, 13), the value of the concentrated records that could be stolen increases by construction. And when coupled with the ACA’s push to reform delivery systems through digitization (viz. through EHRs and health information exchanges (HIEs)), the concern is evident. Because a larger number of patients are being treated within the bounds of hospitals (7), where the encounter typically generates a digital record (initially due to the HITECH Act and further propelled by the ACA), there is more and richer data to steal. When discussing the reach of the ACA and its digitization efforts, Fontenot (14) notes: “With an electronic record, the patient’s entire transition through life and treatment becomes available far beyond that patient and their encounters with the health care system” (p. 74).

On the other hand, this inference does not account for extant knowledge of digitization and the efficacy of statutes designed to limit malfeasance. Numerous scholars have noted that while digitization can result in negative externalities like insurance upcoding, it also streamlines the auditing process, making it easier to detect and correct misconduct (15-17). Similar views have been advanced regarding fraud detection in a digitized healthcare system (i.e., analytical auditing) (14). Second, to the extent that the Health Insurance Portability and Accountability Act of 1996 (HIPAA) mandates rigorous protections of personal health data to limit leakage, data loss might also fall. Insofar as health care systems expanded to accommodate newly-covered patients, it is likely that IT security investments were concomitantly made. If this were the case, the total amount of consumer fraud and identity theft might drop due to the increased data security and oversight of personal health information brought on by the expansion of Medicaid. To explore these competing perspectives, we leverage the phased implementation of Medicaid expansion after the passage of the ACA using a difference in difference approach (18, 19).

## Study Data and Methods

### Data and Sample

To investigate the effect of Medicaid expansion on rates of consumer fraud and identity theft, we construct a unique data set from three sources. First, we gather data on the expansion of Medicaid from the Kaiser Family Foundation. These data are compiled in Appendix Exhibit A1 and indicate the year of Medicaid expansion for each state (if at all). Second, we draw data on claims of fraud and identity theft from the FTC’s Consumer Sentinel Network Reports. These data have been consistently used in empirical research to measure rates of fraud and identity theft (20, 21). We first focus on instances of fraud and identity theft, as opposed to breaches, because there are varying requirements for breach reporting, many breaches go undetected, and breaches vary in the potential harm posed to the public. However, to ensure robustness we also investigate the effect of Medicaid expansion on breached consumer data using the Privacy Rights Clearinghouse (22, 23). Data are organized at the state-year level from 2005 to 2018. Consistent with prior work, we treat the District of Columbia as an independent member of the panel.

### Analytical Approach

To identify the effect, we estimate a two-way fixed effect difference in difference estimation (19). As the dependent variable is a non-negative integer count, we use a Poisson Psuedo Maximum Likelihood (PPML) estimator (24). Formally, we estimate Equation 1, expressed in linear form:

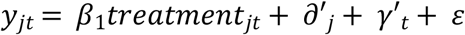

where y_jt_ first takes on the number of identity thefts and then the reported number of cases of consumer fraud. ∂ is the vector of state fixed effects (*j*). γ is the vector of year fixed effects (*t*). ε is the error term. The constant term is not estimated due to the inclusion of the two-way fixed effect structure. Robust standard errors are clustered on the state (i.e. the unit of treatment) (25). Results are in Table 1.

**Table 1:** Effect of Medicaid Expansion on Identity Theft and Fraud.

|  | (1) | (2) | (3) | (4) |
| --- | --- | --- | --- | --- |
| Dependent Variable | PPML<br>Poisson<br>Identity Theft | PPML<br>Poisson<br>Fraud | PPML<br>Poisson<br>Identity Theft | PPML<br>Poisson<br>Fraud |
| Treatment | -0.0747*<br>(0.0448) | -0.177***<br>(0.0599) | -0.0541<br>(0.0774) | -0.102**<br>(0.0470) |
| State Population |  |  | 6.97e-09<br>(1.24e-08) | 4.28e-08***<br>(1.57e-08) |
| Per capita Income |  |  | -3.07e-06<br>(1.55e-05) | -1.69e-05*<br>(9.33e-06) |
| Black Population |  |  | -3.770<br>(3.436) | 4.240<br>(2.983) |
| Latino Population |  |  | 6.97e-09<br>(1.24e-08) | 4.28e-08***<br>(1.57e-08) |
| State FE | Yes | Yes | Yes | Yes |
| Year FE | Yes | Yes | Yes | Yes |
| Observations | 765 | 765 | 343 | 343 |
| Number of Groups | 51 | 51 | 49 | 49 |

Robust standard errors in parentheses (clustered on state). \*\*\* p<0.01, \*\* p<0.05, \* p<0.1

Before discussing our results, several items bear note. The chief concern with any difference in difference estimation is the assessment of pre-treatment trends in the dependent variable (18, 26). The concern is that if heterogeneity exists across the treatment and control groups prior to treatment, we might inappropriately attribute post-treatment difference to the treatment instead of some pre-existing factor. In context, such a concern is not outlandish. If access to healthcare markets influences changing rates of fraud and identity theft, the expansion of Medicaid might be correlated with such factors *a priori*, resulting in a biased estimate, even if the expansion of Medicaid by state legislators as a direct response to general fraud is implausible. To assess this possibility, we estimate a variant of the Autor (27) leads and lags framework, which has become popular in empirical work (28-30). In doing so, we create a series of relative time dummies that capture the temporal distance between treatment in the current period *t* versus treatment in state *j* (conditioned upon the absolute time fixed effects and the location fixed effects). By estimating the effect semi-parametrically, we can visualize the effect over time both pre- and post-treatment. To mitigate power issues in the tails, we collapse all indicators more than 5 years from treatment into a single coefficient. The period immediately prior to treatment is omitted to avoid the dummy variable trap. Results are in Table 2.

**Table 2:** Effect of Medicaid Expansion on Identity Theft and Fraud in Relative Time.

| Estimator | (1) | (2) |
| --- | --- | --- |
| Dependent Variable | PPML<br>Identity Theft | PPML<br>Fraud |
| Rel Time $t_{-5+}$ | 0.197**<br>(0.0853) | 0.183*<br>(0.0984) |
| Rel Time $t_{-4}$ | 0.127**<br>(0.0524) | 0.0894<br>(0.0873) |
| Rel Time $t_{-3}$ | 0.0267<br>(0.0717) | 0.0564<br>(0.0831) |
| Rel Time $t_{-2}$ | -0.125<br>(0.138) | 0.0315<br>(0.0250) |
| Rel Time $t_{-1}$ | -0.00670<br>(0.0561) | 0.0232<br>(0.0226) |
|  | Omitted Period |  |
| Rel Time $t_{+1}$ | -0.00924<br>(0.0652) | -0.149**<br>(0.0601) |
| Rel Time $t_{+2}$ | 0.128***<br>(0.0480) | -0.144***<br>(0.0453) |
| Rel Time $t_{+3}$ | 0.162**<br>(0.0721) | -0.0366<br>(0.0659) |
| Rel Time $t_{+4}$ | 0.0394<br>(0.102) | -0.0653<br>(0.0487) |
| Rel Time $t_{+5+}$ | -0.131<br>(0.129) | 0.00886<br>(0.0650) |
| State FE | Yes | Yes |
| Year FE | Yes | Yes |
| Observations | 765 | 765 |
| Number of Groups | 51 | 51 |

Robust standard errors in parentheses (clustered on state).
\*\*\* p<0.01, \*\* p<0.05, \* p<0.1

### Study Results

In Columns 1-4 in Table 1, we observe a general decrease in rates of identity theft and fraud after Medicaid expansion, although the effects cross beyond the traditional thresholds of significance in Column 3 once controls are added. Still, most columns indicate a negative relationship. Intuitively, this suggests that Medicaid expansion decreases rates of malfeasance, and propose that the mechanisms might be the increased security associated with granting previously uninsured persons access to healthcare markets, increased security through the digitization of these records (a consistent occurrence after the expansion of Medicaid), and/or increased oversight on the treatment of medical records which accords with federal intervention in the market. Economically, estimates indicate a drop in identity theft between ∼5.2% and ∼7.1%. Similarly, estimates indicate a decrease in fraud between ∼9.6% and ∼16.2%.

We turn next to the event study model (Table 2). As can be seen, there is a consistent negative effect of treatment on fraud in Column 2, with negative and significant estimates emerging. Further, as can be seen, there is little in the way of significant pre-treatment trends, although there is a marginally significant difference five years prior to treatment. Further, and consistent with Table 1, in Column 1 we observe no consistent pre- or post-treatment effect, suggesting a *de minimis* relationship between Medicaid expansion and identity theft.

### Data Breaches

To ensure the robustness of the above results it is worth considering alternate measures of data loss. As discussed previously, one popular measure in the empirical literature is to consider the prevalence of reported data breaches and the number of the records stolen (20, 22, 23). While this outcome is more distal from our measures of consumer harm, evaluating the number of breaches offers a valuable check on a potential mechanism, i.e., decreased breaches overall.

To execute these tests, we draw data from the Privacy Rights Clearinghouse, a publicly available repository of historical breaches. We then replicate Equation 1, replacing the number of fraudulent claims and identities stolen with the total number of reported breaches and the total number of breached records. In deference to the number of breaches and the number of records compromised we log the DV and interpret the effect elastically. The estimator is OLS.

Results are in Table 3. As can be seen, the effects are consistent. There is a significant and negative effect on both the number of breaches which have occurred and the total number of compromised records. Two critical takeaways from these estimations are evident. First, these estimations corroborate prior measures and suggests a beneficial effect of Medicaid expansion on data security beyond claims of fraud. Second, and more importantly, they offer suggestive evidence of the mechanism behind the effect, i.e., increased security. To the extent that the rate of breaches and breached records are declining in Medicaid expanding states, it appears that this expansion is slowing data loss by decreasing the number and extent of breaches.

**Table 3:** Effect of Medicaid Expansion on Breaches and Record Theft as Defined by the Privacy Rights Clearinghouse.

| Estimator | (1) | (2) |
| --- | --- | --- |
| Dependent Variable | OLS<br>ln(Breaches) | OLS<br>ln(Records) |
| Treatment | -0.144*<br>(0.0854) | -1.203*<br>(0.613) |
| State FE | Yes | Yes |
| Year FE | Yes | Yes |
| Observations | 765 | 765 |
| R-squared | 0.537 | 0.205 |
| Number of Groups | 51 | 51 |

Robust standard errors in parentheses (clustered on state)
\*\*\* p<0.01, \*\* p<0.05, \* p<0.1

In the interest of space, we refer the interested reader to the Supplemental Appendix for further details on the robustness checks. These include, the implementation of placebo tests, a Goodman-Bacon (31) decomposition, sample truncation, a Callaway and Sant’Anna (32) estimation, and more.

## Discussion

In this work, we investigate the relationship between the expansion of Medicaid provisions under the ACA and data security in the form of fraud and identity theft. The relationship between these concepts is theoretically murky. On the one hand, the injection of hundreds of billions of dollars into the healthcare market materially raises the attractiveness of such targets to malicious actors, notably as this period of time saw an increased rate of personal data digitization (12). As a result, it is plausible that thefts might rise, with the personal information of millions of Americans spilling into secondary markets for the sale of personal data like the dark web (33). On the other hand, there are reasons to believe that cases of identity theft and fraud might fall. Numerous scholars have highlighted the increased security and monitoring which accompanies the digitization of records, and the expansions associated with ACA rollout may have stimulated investments in IT security. Coupled with the fact that patients themselves are now formally covered by insurance, there is an incentive for them to formally engage with the medical system and not misrepresent themselves to physicians.

Upon exploring these competing perspectives, results are three-fold. First, we find that the number of reported cases of fraud significantly declines after the expansion of Medicaid. Second, we find no material or robust change in the number of identities stolen. Third, in deference to traditional approaches to measuring population level data security, we also find a decline in the number of data breaches and stolen or compromised records after the expansion of Medicaid.

Taken in sum, these findings suggest that the mechanism by which the effect manifests is increased oversight, which comes with creating formal markets for underserved communities, and results in increased data security and diminished rates of fraud.

### Policy Implications

While the injection of capital into markets has historically raised the specter of fraud and malfeasance, these concerns appear to be limited when appropriate technological safeguards are implemented. Put simply, risks of moral hazard and fraud have near universally accompanied large infusions of capital into markets (8-11) for obvious reasons. Not only are targets more attractive, because there are more consumers tracked by them (7), but the richness of healthcare data and the interconnected nature of the emergent exchanges has long been thought to lead to data security vulnerabilities (34, 35). Yet, this simplistic view ignores the effect of proactive steps organizations can take (36, 37), and the benefits of direct guidance from federal entities when it comes to safeguarding data (i.e., HIPPA). As a result, to the extent that these organizations appear to be integrating security with organizational and institutional practices (22), they are able to better safeguard consumer data than pre-digitized organizations.

Further, to the extent that any federal program which bluntly injects large amount of capital has been met with skepticism, and raises reasonable concerns of downstream fraud, our findings underscore the importance of coupling capital injection with appropriate controls. Contrast, for example, Medicaid expansion (where tight controls existed) with the Trump Administration’s injections of capital into the economy during the Covid-19 pandemic (e.g., the Paycheck Protection Program (PPP)). To date, the Government Accountability office (38) and NGOs (39) have associated these programs with tens of billions of dollars of fraud against U.S. taxpayers. As a result, numerous federal agencies (ranging from Treasury, to HHS, to the Department of Justice) have been compelled to initiate massive *ex post* efforts to recover these dollars (each of which is also expensive). And while it is beyond the scope of this work to second guess the necessity of capital injections during a global public health emergency, the absence of such controls only bolsters the importance of our findings for policy markets.

Our work also underscores the importance of technological safeguards and regulating inappropriate access. One concern following the digitization of patient records has been that the proliferation of data sharing across organizations could lead to inappropriate use. This is a consistent concern when balancing the size of digital security programs at the local vs. state and federal level. Examples are easy to come by, ranging from the inappropriate search of celebrity health records by medical practitioners (40) to the illegal search of Barack Obama’s records by Philadelphia police officers (41). On the one hand, permitting smaller jurisdictions to manage data is intuitively appealing, because the attractiveness of the target is lower (viz., because fewer records are tracked). However, these entities may not have the resources or technical knowhow to properly safeguard data and limit access, and decentralization may require an unpalatably high ratio of IT investment to overall expenses within each jurisdiction. This once again pushes back on the longstanding assumption that a larger aggregation will lead to an increased likelihood of data loss, notably when aggregation is coupled with superior data controls. We further hope this work serves as a call for future scholarship that investigates the conditions under which greater data aggregation can materially benefit consumer security.

### Limitations

These findings are not without limitation. First, due to the secondary nature of the empirical investigation, we are unable to uncover the exact mechanism by which the decline in fraud manifests. Although our evidence suggests that the number of breaches is declining, it could also be that malicious actors may not be targeting the particular hospitals or physicians’ offices which tend to treat a larger number of Medicaid patients for fear of federal enforcement. It is also possible that formal participation in insurance markets results in fewer cases of fraud on the part of patients (uninsured patients traditionally being without a general practitioner and therefore being less incentivized to engage truthfully with healthcare providers). A third possibility is that physician prevalence for upcoding under digital regimes is declining under Medicaid expansion, again suggesting an enforcement mechanism. We leave further determination of the mechanism to future scholars.

Second, we are reliant on diagnostic methods to ensure the validity of the difference in difference. While the absence of pre-treatment trends and the success of the various falsification tests suggest that the exogeneity assumptions of the DID are met (i.e., treatment can be considered exogenous once conditioned upon controls), this cannot be assured in the absence of laboratory conditions. Bearing this in mind, legislative histories are, to the best of our understanding, devoid of references to data security when state legislatures were debating the expansion of Medicaid. Third, the ACA was expanding coverage in an aggressively changing market, making it difficult to ignore the possibility of an omitted variable bias, despite the successful robustness checks. While the fixed effect structure should minimize the effect of other federal laws (such as the HITECH Act which was implemented universally across the country), the possibility of other state level initiatives remains a possibility.

## Conclusion

This work addresses the possibility that the expansion of Medicaid under the ACA had a material effect on the security of citizen’s personal data. We find no evidence of increased fraud. Instead, we observed rates of fraud declining in Medicaid expanding states, with breaches and breached records declining as well. We hope this work serves as a call to action for researchers on two fronts. First, to consider the effect of Medicaid expansion on broader social issues besides healthcare. While some scholars are beginning to take this approach (3, 4, 42), it remains an underserved area of research for one of the largest federal programs in U.S. history. Second, we hope this work pushes scholars to break the mold of focusing solely on data breaches in favor of investigating instances of fraud and identity theft (i.e., materialized harm to consumers). While the focus on breaches is appealing, it is potentially problematic because breach discovery is not 100%, and not all breaches result in material harm, which can result in inconsistent reporting. Pivoting away from this approach, and towards observable instances of harm, offers the opportunity for researchers to begin resolving this discrepancy between measures and constructs.

## Data Availability

Data are available via GitHub please contact the authors for details.

